# Sex-differences in determinants of suicide risk preceding psychiatric admission: An electronic medical record study

**DOI:** 10.1101/2020.11.19.20227694

**Authors:** Robyn J. McQuaid, Katerina Nikolitch, Katie L. Vandeloo, Patricia Burhunduli, Jennifer L. Phillips

## Abstract

Using electronic medical record (EMR) data collected from psychiatric inpatient admissions, the objective of this study was to identify sex differences in risk factors for presence of suicide plans and/or attempts within the 30 days preceding hospital admission. Resident Assessment Instrument for Mental Health (RAI-MH) intake data were obtained for patients admitted to a Canadian tertiary-care hospital deemed a ‘threat or danger to self’ during a ten-year period (2008-2018). Data was extracted for individuals categorized into three groups: non-suicidal (N=568), presence of suicide plan (N=178), and presence of suspected suicide attempt (N=124) in the 30 days prior to hospital admission. A multivariate logistic regression revealed that younger age (odds ratio=0.97), female sex (OR=1.56), disrupted family relationships (OR=1.54), recent stressors (OR=1.59), participation in social activities (OR=1.54), having no confidant (OR=1.55), and diagnosis of depressive disorder (OR=5.54) increased the odds of suicide plan and/or attempt in the 30 days prior to hospital admission. Stratifying the regression model by sex highlighted different risk factors for suicide plan and attempt specific to males and females. EMR-derived findings highlight psychosocial and clinical determinants associated with suicide plan or attempt prior to psychiatric admission that differ according to sex.

## 1. Introduction

Approximately 4,000 Canadians die by suicide every year^1^ and for every suicide death there are an estimated 20 suicide attempts.^2^ Suicide disproportionately affects specific populations and groups,^3-7^ yet over 90% of individuals who die by suicide meet criteria for at least one psychiatric diagnosis and 60% meet criteria for a mood disorder.^8,9^ Within psychiatric populations, at highest suicide risk are individuals with severe mental illness that necessitates inpatient care.^10^ Psychiatric inpatient admission often occurs at times of crisis when individuals might pose a threat of harm to themselves or others,^11^ however even within this population, identifying those at highest risk of suicide remains difficult. An additional challenge is to identify precipitating factors that may elevate imminent risk of suicide in vulnerable individuals.

Stressful life circumstances such as loss of income and residential instability have been associated with increased risk of suicidal ideation (thoughts of killing oneself which may include a suicide plan) and suicide attempts (self-injurious behaviour accompanied by an intent to die).^12-14^ One’s social climate preceding the onset of suicidal behaviours is also important, and perceived loneliness uniquely predicts suicide attempt above and beyond depression.^15^ Furthermore, targeting hopelessness (a common symptom of major depressive disorder) appears to significantly reduce the depression-suicide link in both males and females.^16^ Identifying demographic and psychosocial factors present at the time of elevated suicide risk may lead to improved recognition of suicide vulnerability; unfortunately, existing literature on this subject remains sparse.

Nationally reported statistics on suicide rates distinguish discrepancies between males and females. Females appear three-to-four times more likely to attempt suicide than males,^17,18^ and are admitted to hospital following suicide attempt or self-injury more frequently.^19,20^ However, males are three times more likely to die by suicide.^17,21^ Such differences are often attributed to the fact that depression (a known risk factor for suicide) is more prevalent in females^22^ and that males tend to use more lethal means accounting for their higher suicide death rates.^23^ Moreover, among females with a previous suicide attempt, severity of self-reported depressive symptoms predicted subsequent suicide attempts, an effect not observed in males.^24^ The potential contributing role of psychosocial factors on sex-specific differences in suicidal behaviour requires further study.^25^

Recent research suggests that suicide risk evaluation could be improved through simultaneous use of electronic medical record (EMR) data and standardized risk assessment tools.^26^ Since 2008 in the Province of Ontario, all psychiatric inpatient service providers are required to assess patients at admission, discharge, and quarterly in the case of extended stays using the Resident Assessment Instrument for Mental Health (RAI-MH) (http://www.interrai.org/).^27^ RAI-MH admission data provides a snapshot of the demographic, psychosocial and clinical circumstances of individuals within an acute period prior to their hospital admission. Harnessing this data may permit identification of sex-specific factors present at the time of elevated suicide risk in individuals who had suicide plans or attempts in the period immediately preceding hospitalization.

The objectives of the current investigation were to: 1) identify factors associated with elevated suicide risk in the 30 days prior to admission; 2) identify factors that differentiate individuals with a suicide plan from those with a suspected suicide attempt; 3) determine whether suicide risk factors differ between males and females; and 4) compare mental health symptoms at the time of admission in relation to suicide risk.

## 2. Methods

### 2.1. Data source

This study was a secondary analysis of EMR data recorded by clinicians at the point of psychiatric admission accessed from the RAI-MH database at the Royal Ottawa Mental Health Centre (ROMHC) in Ottawa, Ontario, Canada. De-identified RAI-MH data were obtained for all admission cases in which “threat or danger to self” was coded as one of the “reasons for admission” over a ten-year period, from January 1, 2008 to December 31, 2017. Data from all ROMHC inpatient units (crisis, forensic, geriatric, mood, recovery, schizophrenia, and youth) were obtained. For individuals with multiple admissions over the ten-year period, only data from first admissions was included in the current study to avoid redundancy. From the full dataset, select admission cases were included according to operationally defined groups based on pre-admission suicide risk level (see **Figure 1**). This study received approval from the ROMHC Research Ethics Board.

**Figure 1.**
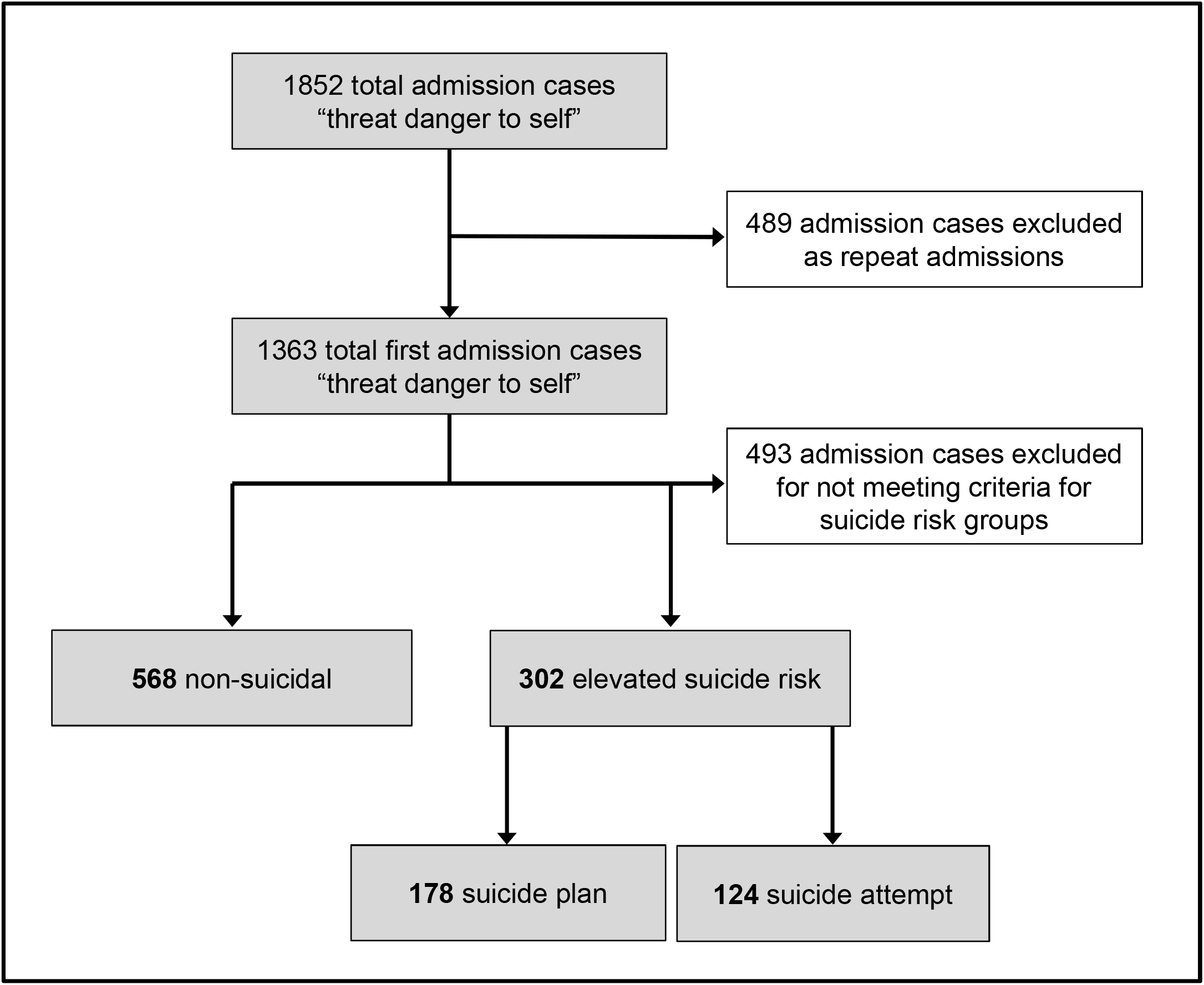
A flow diagram depicting the sample criteria and exclusions.

### 2.2. Suicide groups

Individuals were organized into three groups based on self-injury related variables: 1) **Non-suicidal**, operationally defined as admission case records where the data elements “considered performing a self-injurious act” and “most recent self-injurious act” were not endorsed for the previous 1 year period, there was no suicide plan in the 30 days prior to hospital admission, and no lifetime history of fatal-intent self-injurious attempt; 2) **Suicide plan**, defined as endorsement of the presence of a suicide plan in the 30 days preceding admission but no “self-injurious attempt” within that time period; and 3) **Suspected suicide attempt**, defined as coding of “most recent self-injurious attempt” within the 30 days prior to hospital admission, and positive endorsement of “intent of any self-injurious attempt was to kill himself/herself”. As the RAI-MH does not directly measure suicide attempts, these two variables were combined to reduce inclusion of individuals with recent engagement in non-suicidal self-injury in the suicide attempt group.

### 2.3. Patient-level variables

The RAI-MH provides more than 300 individual data elements. Selecting plausible suicide risk variables based on previous literature and focusing on the time period immediately preceding admission, we extracted the following patient-level variables for each admission case: sex, age, marital status, education, sources of income, residential stability, stressors/life events, family roles, social relations, participation in social activities of long-standing interest, and psychiatric diagnosis. Certain variables were dichotomized to binary variables including education (“incomplete high school” yes/no) and marital status (“single/unpartnered” yes/no). For stressors, the RAI-MH lists individual variables for 16 adverse life events. Stressor categories were pooled into a single variable and binary coded as having occurred within the 30 days preceding admission (“stressor present” yes/no). For family roles, positive endorsements of the RAI-MH variable “belief that relationship(s) with immediate family members is disturbed or dysfunctional” by the patient, family/friends, or both were pooled and the variable was binary coded (“belief present” yes/no). Participation in social activities was binary coded according to occurrence during the 30 days preceding admission. Psychiatric diagnoses were determined by a physician during initial inpatient assessment using Diagnostic and Statistical Manual of Mental Disorders (DSM) criteria.^28,29^ The RAI-MH also includes mental state indicators (scored as continuous variables) that report psychiatric symptom severity during the first 3 days of psychiatric hospitalization. 13 mental state indicators were selected for examination.

### 2.4. Statistical analysis

Data were analyzed with IBM SPSS Statistics (v26). A multivariate logistic regression model was conducted to investigate potential risk factors predicting the presence of suicide plan or attempt in the 30 days preceding hospital admission. For this analysis, individuals with either a suicide plan or suspected attempt were combined and compared to non-suicidal individuals. Additionally, a multivariate logistic regression model was conducted to differentiate individuals with a suicide plan from those with a suicide attempt (non-suicidal individuals were excluded from this analysis). To investigate the association between predictor variables and suicide risk specifically in males and females, the logistic regression models described above were conducted stratified by sex. Finally, multivariate ANOVAs were performed to compare mental state indicators for individuals deemed as non-suicidal versus those at elevated suicide risk (with suicide plan or attempt).

## 3. Results

### 3.1 Descriptive variables

A total of 1,852 admission cases were provided by the ROMHC. The final dataset comprised admission cases from 870 individuals, *M*_*age*_ = 48.3 years (*SE* = .75, range 15-97 years), of which 462 were females (53.1%) and 408 were males (46.9%). At the time of admission, 36.4% of individuals had not completed high school, and 60.3% reported residential instability. Moreover, 9.0% did not have a source of income; and, of those receiving income, only 6.3% was derived from employment. Remaining income sources included pension (45.9%), disability insurance (21.0%) and social assistance (11.0%). Demographic information is displayed according to groups in **Table 1**.

**Table 1.** Descriptives of non-suicidal and elevated suicide risk groups.

| Characteristics | Non-Suicidal<br>(N=568) | Suicide Plan<br>(N=178) | Suicide Attempt<br>(N=124) | Elevated<br>Suicide Risk<sup>a</sup><br>(N=302) |
| --- | --- | --- | --- | --- |
| Age, years (M, SD) | 52.3 ($\pm 0.90$) | 41.6 ($\pm 1.46$) | 39.2 ($\pm 1.75$) | 40.6 ($\pm 1.23$) |
| Sex |  |  |  |  |
| Male | 57.2% | 43.3% | 48.4% | 45.4% |
| Female | 42.8% | 56.7% | 51.6% | 54.6% |
| No source of income | 5.6% | 17.4% | 12.1% | 15.2% |
| Did not complete<br>highschool | 35.1% | 37.9% | 40.3% | 38.9% |
| Residential instability | 62.1% | 61.2% | 50.8% | 57.0% |

<sup>a</sup>The elevated suicide risk group combines the suicide plan and suicide attempt groups

Psychosocial and clinical factors were also assessed that were deemed relevant to suicide plans and attempts. At the time of admission, 18.4% of individuals reported having no confidant and 24.6% had disturbed or dysfunctional relationships with immediate family members. 24.9% of individuals experienced a stressor in the 30 days preceding admission. Reported stressors were similar for males and females with the most common being review hearing (7.0%), distress about the health of another person (6.6%), and conflict-laden relationship (6.1%). 44.7% of individuals had not participated in social activities of long-standing interest in the 30 days preceding admission. According to DSM diagnostic categories, included individuals had primary diagnoses of schizophrenia or other psychotic disorder (41.0%), depressive disorder (24.8%), neurocognitive disorder (14.6%), bipolar disorder (9.5%), substance-related or addictive disorder (3.5%), trauma or stressor-related disorder (2.8%), or other categories (each < 1.0%).

### 3.2. Logistic regressions

#### 3.2.1. Model assessing the risk factors of elevated suicide risk

A logistic regression model comprising the main hypothesized variables was conducted to identify predictors of elevated suicide risk (presence of suicide plan or attempt) in the 30 days preceding admission. The overall model was significant (χ^2^ (10)= 182.09, *p*<.001, R^2^ =.21). Increased odds of elevated suicide risk was associated with younger age, (OR=0.97, 95% CI=0.97-0.98, *p*<.001), female sex, (OR=1.56, 95% CI=1.01-2.21, *p*=.01), disrupted family relationships, (OR=1.54, 95% CI=1.04-2.28, *p*=.03), presence of a recent stressor, (OR=1.59, 95% CI=1.10-2.31, *p*=.01), continued participation in social activities, (OR=1.54, 95% CI=1.09-2.19, *p*=.02), having no confidant, (OR=1.55, 95% CI=1.02-2.37, *p*=.04), and diagnosis of a depressive disorder (OR=5.54, 95% CI=3.71-8.27, *p*<.001). Sources of income (*p*=.24), education (*p*=.25), and residential instability (*p*=.31) were not significant in this model.

#### 3.2.2. Sex-based analyses assessing risk factors of elevated suicide risk

The overall model was stratified by sex to examine specific predictors of elevated suicide risk for males and females. This model was significant for males (χ^2^ (9)=48.89, *p*<.001, R^2^ =.11). The predictors that significantly increased the odds of suicide plan/ attempt for males were having no confidant (OR=2.13, 95% CI=1.19-3.80, *p*=.01), presence of recent stressors (OR=1.95, 95% CI=1.16-3.29, *p*=.01), continued participation in social activities (OR=1.67, 95% CI=1.02-2.71, *p*=.04), and depressive disorder diagnosis (OR=3.89, 95% CI=2.14-7.06, *p*<.001). All other variables including age (*p*=.28), education (*p*=.25), sources of income (*p*=.23), residential instability (*p*=.39), and disrupted relationships (*p*=.23) were non-significant.

For females, the overall model was also significant (χ^2^ (9) =140.42, *p*<.001, R^2^ = .31), and predictors associated with increased odds of suicide plan/attempt included younger age (OR=0.96, 95% CI=0.94-0.97, *p*<.001), and diagnosis of a depressive disorder (OR=7.04, 95% CI=3.97-12.47, *p*<.001). None of the other predictors were significant including sources of income (*p*=.63), education (*p*=.77), residential instability (*p*=.73), social activities (*p*=.26), disrupted relationships (*p*=.15), having no confidant (*p*=.67), or recent stressors (*p*=.31).

#### 3.2.3. Model assessing risk factors for suicide plan versus attempts

The overall model assessing risk factors to differentially identify individuals who had a suicide plan versus those who had an attempt in the 30 days preceding hospital admission was not significant (χ^2^ (10) =10.34, *p*=.28, R^2^ =.04).

### 3.3. Mental state indicators associated with elevated suicide risk

A multivariate ANOVA was conducted to distinguish mental state indicators in individuals with a recent suicide plan or attempt from those deemed non-suicidal in the period preceding admission. The overall model was significant (*Pillai’s Trace* =. η^2^ 20, *F* (14, 855)=15.40, *p*<.001, η^2^*=*.20). As shown in **Table 2**, a number of indicators differed according to suicide risk, including: presence of sad, pained, worried facial expressions (*p*=0.001, η^2^*=* .01), making negative statements (*p*<.001, η^2^*=*.07), self-deprecation (*p*<.001, η^2^ *=*.08), expressions of guilt or shame (*p*<.001, η^2^ *=*.09), statements of hopelessness (*p*<.001, η^2^= .11), irritability (*p*<.001, η^2^*=*.03), anxious complaints (*p*<.001, η^2^*=*.03, η^2^ *=*.04), anhedonia (*p*<.001, η^2^ *=*.04), loss of interest (*p*<.001, η^2^ *=*.02), lack of motivation (*p*=.001, η^2^*=*.01), and sleep problems (*p*=.026, η^2^*=*.01). However, two mental state indicators, episodes of panic (*p*<.10), and command hallucinations (*p*=.21), were not significant.

**Table 2.** Mental state indicators of non-suicidal and elevated suicide risk groups.

| Clinical Features | Non-Suicidal<br>(M±SE) | Elevated suicide risk<sup>a</sup><br>(M±SE) |
| --- | --- | --- |
| Facial Expression** | 1.77 ± 0.05 | 2.09 ± 0.07 |
| Made Negative Statements*** | 1.09 ± 0.05 | 1.81 ± 0.07 |
| Self-Deprecation*** | 0.53 ± 0.05 | 1.20 ± 0.06 |
| Guilt/Shame*** | 0.44 ± 0.04 | 1.10 ± 0.06 |
| Hopelessness*** | 0.54 ± 0.05 | 1.34 ± 0.06 |
| Irritability*** | 1.47 ± 0.05 | 0.99 ± 0.07 |
| Anxious Complaints*** | 1.25 ± 0.05 | 1.80 ± 0.07 |
| Episodes of Panic | 0.37 ± 0.03 | 0.29 ± 0.05 |
| Anhedonia*** | 0.77 ± 0.05 | 1.31 ± 0.07 |
| Loss of Interest*** | 1.08 ± 0.05 | 1.48 ± 0.08 |
| Lack of Motivation** | 1.21 ± 0.06 | 1.53 ± 0.08 |
| Command Hallucinations | 0.29 ± 0.03 | 0.35 ± 0.07 |
| Sleep Problems* | 0.96 ± 0.05 | 1.16 ± 0.07 |

\* $p < 0.05$ , \*\* $p < 0.01$ , \*\*\* $p < 0.001$
<sup>a</sup>The elevated suicide risk group combines the suicide plan and suicide attempt groups

#### 3.3.1. Sex-based analyses assessing mental state indicators related to elevated suicide risk

Importantly, the multivariate ANOVA was significant when examining males alone (*Pillai’s Trace* = .22, *F* (13, 448)=9.86, *p*<.001, η^2^*=*.22). While results remained similar to the above model with both males and females, in this model, sleep problems became non-significant (*p*=.23), in addition to episodes of panic (*p*=.39) and command hallucinations (*p*=.26). This model was also significant among females (*Pillai’s Trace* =.18, *F* (13, 394)=7.04, *p*<.001, η^2^*=* .18). For females, results were similar to males in that sleep problems (*p*=.09) and command hallucinations (*p*=.36) were non-significant. However, unlike males in which sad, pained, worried facial expression was significant (*p*<.01), facial expression was not significant for females (*p*=.12), while episodes of panic was significant (*p*<.001, η^2^*=*.03).

#### 3.3.2. Mental state indicators associated with suicide plan versus attempts

Upon comparing mental state indicators in those with a suicide plan to those with an attempt prior to hospital admission, the overall model was not significant (*Pillai’s Trace* =.85, *F* (13, 288)=1.15, *p*=.32, η^2^*=*.05).

## 4. Discussion

The current study harnessed EMR data from individuals admitted to a Canadian tertiary care psychiatric hospital deemed at risk to themselves. More than half of the sample experienced residential instability at the time of admission, which is consistent with research highlighting the strong connection between instable housing and psychiatric disorders.^30^ Moreover, few individuals received income through employment, with the majority receiving a pension, social assistance or disability insurance. These basic demographic data provide support to literature linking indicators of low socioeconomic (SES) status as risk factors for mental illnesses.^31-33^ However, despite high rates of residential instability and low employment, SES indicators were not significant predicators of suicide plans/attempts, possibly due to the SES homogeneity of the sample.

Diagnosis of depression was the strongest predictor of suicide plans/attempts across all models. Indeed, individuals with a primary depressive disorder diagnosis were at least five times more likely to have a suicide plan or attempt. This is in-line with findings that 60% of individuals who die by suicide meet criteria for a mood disorder.^8,9^ A diagnosis of depression is also a significant longitudinal predictor for suicidal ideation and attempts, although a recent meta-analysis suggests the effects of depression on suicide outcomes, including ideation, attempts and deaths, are not as strong as expected.^34^ Examining specific features of depression in relation to suicide outcomes might prove beneficial in identifying individual risk. In this regard, when considering hopelessness, the depression-suicide relationship is significantly reduced,^16^ and hopelessness is associated with suicidal ideation independent of depression severity.^35^ The current findings reveal that hopelessness had the largest effect size in relation to recent suicide plan/attempts across all mental state indicators, highlighting the potential importance of early identification of hopelessness. Expressions of guilt and shame had the next largest effect on suicide risk. In a recent meta-analysis, guilt and shame were associated with non-suicidal self-harm, however, few reports have examined these negative emotions in relation to suicide outcomes.^36,37^ Thus, the current findings suggest that specific clinical symptoms that are closely tied to depression, such as hopelessness, guilt and shame may help identify elevated suicide risk in an inpatient psychiatry population.

Across males and females, a depressive disorder diagnosis was a significant and strong predictor for suicide plans and attempts, however, among females, depression increased the risk of suicide plans/attempts seven times, compared to just under four times for males. Based on these data it was not surprising that female sex was a significant predictor of elevated suicide risk. Our data supports findings that females are more likely to attempt suicide compared to males.^17,18^ However, while males are more likely to die by suicide,^17, 21^ the gap between males and females is narrowing as the suicide rate for females in the U.S. increased 50% from 2000-2016, compared to a 21% increase for males during this time.^38^ Moreover, the proportion of individuals with past-year suicidal ideation who attempted suicide from 2008-2017 was significantly higher among females in the U.S. and those aged 18-25 years.^39^ In Canada, females are more frequently admitted to hospital following suicide attempt or self-injury.^19,20^ Of particular concern is the alarming recent increase in young females hospitalized due to self-injury in Canada. From 2009 to 2014 the rate of intentional self-harm related hospitalizations increased by 110% in females and 35% for males aged 10-17 years.^40^ Similarly, from 2003-2017 increased rates of Ontario emergency department visits for self-harm and mental health were found for all youth aged 13-17 years, but especially among females.^41^ Herein we report that both female sex and younger age were associated with elevated suicide risk, supporting these emerging trends.

For males, a number of psychosocial factors, including having no confidant, participating in social activities and presence of recent stressors were associated with increased risk of suicide plan or attempt. As stress is a longitudinal predictor of both suicidal ideation and attempts,^42,43^ it was expected that stressful events would be identified as a significant risk factor. Males were also twice as likely to report recent suicide plans or attempts if they had no confidant. Due to masculinity norms, such as self-reliance, men are less likely to seek help for mental health issues.^44^ Although men express wanting to talk about their mental health and developing relationships to support these discussions,^45^ as in the current study, having no confidant would serve as a barrier. Surprisingly, although similar proportions of males and females reported not having a confidant, it was not a significant risk factor for females. However, because depression was such a strong risk factor for females (OR=7.04), it is possible that it obfuscated the effect of other variables. For males, the finding that participation in social activities of long-standing interest was positively associated with suicide plans/attempts was unexpected. However, these interactions may not have been positive, in-line with data showing that expecting support and instead experiencing unsupportive social interactions is strongly related to poor mental health.^46^ It is also possible that males with suicidal ideation made an effort to disguise their worsening mental state by engaging in their usual associations and activities. This is consistent with reports that males are more likely than females to self-stigmatize depression and suicidal thoughts and behaviours.^47^ In any case, our results show that psychosocial factors such as stressful experiences and social relationships are particularly important in screening for elevated suicide risk among males.

This study used EMR data to examine demographic and psychosocial characteristics of individuals at elevated suicide risk prior to psychiatric hospital admission. Strengths of the study included the large, representative sample of psychiatric inpatients and careful examination of sex differences. The study also had a number of limitations. First, the RAI-MH does not directly measure suicide attempts, thus, suspected suicide attempts were derived from two separate variables. Ultimately, it would have been preferable to have suicide attempts confirmed and coded in the data. Second, the study comprised data from a single tertiary care psychiatric hospital with no emergency department. While this could also be viewed as a strength, as this investigation was targeted toward inpatients with severe mental illness who are deemed at high risk of suicide, it is also a limitation as there could be delays between first presenting at another hospital with an emergency department and admission to the ROMHC. Third, data was obtained only for individuals deemed a “threat or danger to self” at admission. Those we classified as nonsuicidal (absence of recent history of suicidal ideation, plans or self-injury) may have had greater issues with self-care or specific psychiatric symptoms that posed a threat to their well being rather than being at risk of suicide at admission. Fourth, while a number of demographic data were included, data on race/ethnicity and gender identity are not fully collected in the RAI-MH. Thus, while we recognize that suicide disproportionately affects specific and/or marginalized populations,^3-7^ we were not able to examine these factors in our models.

Predicting suicide on an individual level remains exceptionally challenging. Ultimately, we could not tease apart those who transition from suicide plan to attempt. To help differentiate the transition from suicide thoughts to behaviours, it might be beneficial to develop new approaches to understanding suicide, such as the use of artificial intelligence (AI),^48^ which might prove valuable in predicting suicide risk beyond hypothesis driven approaches. Despite this, the current findings highlight risk factors associated with progression towards forming a suicide plan or attempt. Thus, identifying individuals at highest risk of suicide, particularly in a high-risk population, is an important first step towards suicide prevention efforts. Together, our findings suggest that depression is a very strong risk factor for suicide plans and attempts, particularly for females and it highlights that younger females in particular might be at elevated suicide risk. Finally, these data reveal that for males, having no confidant doubles the risk of suicide plans/attempts, suggesting that interventions to provide social support to men in distress and increase help seeking behaviours for mental health could prove beneficial for suicide prevention.

### Data Access Statement

The current investigation is a secondary analysis of electronic medical record data from the Royal Ottawa Mental Health Centre. Thus, this data is not owned by the authors and cannot be made publically available. However, this and other Ontario hospital data collected through the Ontario Mental Health Reporting System (OMHRS) can be requested from the Canadian Institutes of Health Information.

## Acknowledgements

The authors thank the information technology staff at the Royal Ottawa Mental Health Centre for compiling the data.

## References

1. Statistics Canada. Leading causes of death, total population, by age group. 2018. https://www150.statcan.gc.ca/t1/tbl1/en/tv.action?pid=1310039401&pickMembers%5B0%5D=2.1&pickMembers%5B1%5D=3.1. Accessed August 20, 2020.

2. World Health Organization. Suicide data. 2016. https://www.who.int/mental_health/prevention/suicide/suicideprevent/en/. Accessed April 18, 2016.

3. Kirmayer LJ, Brass GM, Holton T, et al. Suicide among Aboriginal People in Canada (PDF), Ottawa, Ontario: The Aboriginal Healing Foundation. 2007. http://www.ahf.ca/downloads/suicide.pdf. Accessed November 17, 2018.

4. McQuaid RJ, Bombay A, McInnis OA, et al. Suicide ideation and attempts among First Nations peoples living on-reserve in Canada: The intergenerational and cumulative effects of Indian Residential Schools. Can J Psychiatry. 2017;62(6):422–430.

5. Peter T, Edkins T, Watson R, et al. Trends in suicidality among sexual minority and heterosexual students in a Canadian population-based cohort study. Psychol Sex Orientat Gend Divers. 2017;4(1):115–123.

6. Sapers H. A three year review of Federal inmate suicides (2011-2014). 2014. https://www.oci-bec.gc.ca/cnt/rpt/oth-aut/oth-aut20140910-eng.aspx. Accessed 26 August, 2020).

7. Sareen J, Afifi TO, Taillieu T, et al. Trends in suicidal behaviour and use of mental health services in Canadian military and civilian populations. CMAJ. 2016;188(11):E261–267.

8. Arsenault-Lapierre G, Kim C, Turecki G. Psychiatric diagnoses in 3275 suicides: A meta-analysis. BMC Psychiatry. 2004;4(37):1–11.

9. Cavanagh JTO, Carson AJ, Sharpe M, et al. Psychological autopsy studies of suicide: a systematic review. Psychol Med. 2003;33(3):395–405.

10. Hjorthøj CR, Madsen T, Agerbo E, et al. Risk of suicide according to level of psychiatric treatment: a nationwide nested case-control study. Soc Psychiatry Psychiatr Epidemiol. 2014;49(9):1357–1365.

11. Ziegenbein M, Anreis C, Brüggen B, et al. Possible criteria for inpatient psychiatric admissions: which patients are transferred from emergency services to inpatient psychiatric treatment? BMC Health Serv Res. 2006;6:150.

12. Cupina, D. Life events, gender and suicidal behaviours in the acute community setting. Australas Psychiatry. 2009;17(3):233–236.

13. Pitman A, Krysinska K, Osborn D, et al. Suicide in young men. Lancet. 2012;379(9834):2383–2392.

14. Skopp NA, Zhang Y, Smolenski DJ, et al. Risk factors for self-directed violence in US soldiers: a case-control study. Psychiatry Res. 2016;245:194–199.

15. Levi Y, Horesh N, Fischel T, et al. Mental pain and its communication in medically serious suicide attempts: an “impossible situation”. J Affect Disord. 2008;111(2-3): -244-250.

16. Zhang J, Li Z. The association between depression and suicide when hopelessness is controlled for. Compr Psychiatry. 2013;54(7):790–796.

17. Bachmann, S. Epidemiology of suicide and the psychiatric perspective. Int J Environ Res. 2018;15(7):1425.

18. Chang B, Gitlin D, Patel R. The depressed patient and suicidal patient in the emergency department: evidence-based management and treatment strategies. Emerg Med Pract. 2011;13(9):1–23.

19. Langlois S, Morrison P. Suicide deaths and suicide attempts. Health Reports. 2002;13(2):9–21.

20. Skinner R, McFaull S, Draca J, et al. Suicide and self-inflicted injury hospitalizations in Canada (1979 to 2014/15). HPCDP. 2016;36(11):243–251.

21. GBD 2015 Mortality and Cause of Death Collaborators. Global, regional, and national life expectancy, all-cause mortality, and cause-specific mortality for 249 causes of death, 1980–2015: a systematic analysis for the Global Burden of Disease Study 2015. Lancet. 2016;388(10053):1459–1544.

22. Kuehner C. Gender differences in unipolar depression: an update of epidemiological findings and possible explanations. Acta Psychiatr Scand. 2003;108:163–174.

23. Mergl R, Koburger N, Heinrichs K, et al. What are reasons for the large gender differences in the lethality of suicidal acts? An epidemiological analysis in four European countries. PLOS One. 2015;10(7):e0129062.

24. Monnin J, Thiemard E, Vandel P, et al. Sociodemographic and psychopathological risk factors in repeated suicide attempts: gender differences in a prospective study. J Affect Disord. 2012;136(1-2):35–43.

25. Vijayakumar L. Suicide in women (Suppl 2). Indian J Psychiatry. 2015;57(6):S233–S238.

26. Bruer RA, Rodway-Norman M, Large M. Closer to the truth: admission to multiple psychiatric facilities and an inaccurate history of hospitalization are strongly associated with inpatient suicide. Can J Psychiat. 2018;63(11):748–756.

27. Hirdes JP, Smith TF, Rabinowitz T, et al. The Resident Assessment Instrument–Mental Health (RAI-MH): inter-rater reliability and convergent validity. J Behav Health Serv Res. 2002;29(4):419–432.

28. American Psychiatric Association: Diagnostic and Statistical Manual of Mental Disorders, 4th ed, text revision (DSM-IV-TR). Washington, DC, American Psychiatric Association, 2000.

29. American Psychiatric Association: Diagnostic and Statistical Manual of Mental Disorders, 5th ed (DSM-V). Washington, DC, American Psychiatric Association, 2013.

30. Fazel S, Khosla V, Doll H, et al. The prevalence of mental disorders among the homeless in western countries: systematic review and meta-regression analysis. PLoS Med 2008;5(12):e225.

31. Kosidou K, Dalman C, Lundberg M, et al. Socioeconomic status and risk of psychological distress and depression in the Stockholm Public Health Cohort: a population-based study. J Affect Disord. 2011;134(1-3):160–167.

32. Lund C, Breen A, Flisher AJ, et al. Poverty and common mental disorders in low and middle income countries: A systematic review. Soc Sci Med. 2010;71(3):517–528.

33. McLaughlin KA, Costello EJ, Leblanc W, et al. Socioeconomic status and adolescent mental disorders. Am J Public Health. 2012;102(9):1742–1750.

34. Ribeiro JD, Huang X, Fox KR, et al. Depression and hopelessness as risk factors for suicide ideation, attempts and death: meta-analysis of longitudinal studies. Br J Psychiatry. 2018;212(5):279–286.

35. Wolfe KL, Nakonezny PA, Owen VJ, et al. Hopelessness as a predictor of suicide ideation in depressed male and female adolescent youth. Suicide Life Threat Behav. 2019;49(1):253–263.

36. Sheehy K, Noureen A, Khaliq A, et al. An examination of the relationship between shame, guilt and self-harm: A systematic review and meta-analysis. Clin Psychol Rev. 2019;73:101779.

37. Bryan CJ, Morrow CE, Etienne N, et al. Guilt, shame, and suicidal ideation in a military outpatient clinical sample. Depress Anxiety. 2013;30(1):55–60.

38. Hedegaard H, Curtin SD, Warner M. Suicide rates in the United States continue to increase. NCHS Data Brief. 2018;309:1–8.

39. Lange S, Bagge C, Probst C, et al. Proportion of individuals with past-year suicidal ideation who attempted suicide over the past 10 years in the United States, and the influence of age and sex. Crisis. 2020;Eub ahead of print, May 19, 2020. doi: 10.1027/0227-5910/a000690.

40. Canadian Institutes for Health Information. Intentional self harm among youth in Canada. 2014. https://www.cihi.ca/sites/default/files/info_child_harm_en.pdf. Accessed August 20, 2020.

41. Gardner W, Pajer K, Cloutier P, et al. Changing rates of self-harm and mental disorders by sex in youths presenting to Ontario emergency departments: repeated cross-sectional study. Can J Psychiatry. 2019;64(11):789–797.

42. Chen YL, Kuo PH. Effects of perceived stress and resilience on suicidal behaviors in early adolescents. Eur Child Adolesc Psychiatry. 2020;29(6):861–870.

43. Park CHK, Lee JW, Lee SY, et al. Characteristics of the “young-old” and “old-old” community-dwelling suicidal ideators: a longitudinal 6-month follow-up study. Compr Psychiatry. 2019;89:67–77.

44. Wong YJ, Ho MHR, Wang SY, et al. Meta-analyses of the relationship between conformity to masculine norms and mental health-related outcomes. J Couns Psychol. 2017;64(1):80–93.

45. Herron RV, Ahmadu M, Allan JA, et al. “Talk about it:” changing masculinities and mental health in rural places? Soc Sci Med. 2020;258:113099.

46. Woods R, McInnis O, Bedard M, et al. Social support and unsupportive interactions in relation to depressive symptoms: implication of gender and the BDNF polymorphism. Soc Neurosci. 2020;15(1):64–73.

47. Oliffe JL, Ogrodniczuk JS, Gordon SJ, et al. Stigma in male depression and suicide: a Canadian sex comparison study. Community Ment Health J. 2016;52(3):302–310.

48. Gradus JL, Rosellini AJ, Horváth-Puhó E, et al. Prediction of sex-specific suicide risk using machine learning and single-payer health care registry data from Denmark. JAMA Psychiatry. 2020;77(1):25–34.

